# Signals reported to Rucaparib: An Updated Comprehensive Disproportionality Analysis Using FDA Adverse Event Reporting System

**DOI:** 10.1101/2024.09.20.24314057

**Authors:** Anshaj Sharma, Pramod Kumar Adusumilli

## Abstract

Ovarian cancer currently ranks as the fourth most common cancer. This cancer is metastatic in nature and making it more challenging to manage. Prostate cancer is the one of the major causes of disease and death among men. Approximately 1.6 million men are diagnosed with prostate cancer. Rucaparib, a poly-ADP ribose polymerase inhibitor has been approved by United States Food and Drug Administration for management of ovarian cancer and prostate cancer. Rucaparib was approved by US Food and Drug Administration in the year 2016 for the treatment of ovarian cancer and 2020 for the treatment of prostate cancer. To assess the potential association between rucaparib and the identified signals, a disproportionality analysis of spontaneous reports is being conducted. Reports were taken from FAERS data base and retrospective case/non case study was conducted. Reporting Odds ratio (ROR), Relative Reporting Ratio (RRR), Chi Squared Value (χ^2^) and Proportional Reporting Ratio (PRR) and Drug Event (DE) were used to perform disproportionality analysis. 144 signals were considered as positive adverse drug reactions using the criteria χ^2^ >4, PRR >2, ROR >2 and DE ≥ 3. Through disproportionality analysis of the FAERS data, signal was identified between the signals-increased prostate specific antigen, decreased serum magnesium levels, decreased glomerular filtration rates, blood iron decreased and vitamin d decreased and rucaparib. The current investigation indicated that rucaparib may increase the incidence of the identified signals.

## Introduction

A drug is a chemical substance used for the treatment, prevention, cure or diagnosis of a disease or a condition in animals or human beings. Drugs approved by USFDA (United States Food and Drug administration) must be effective and safe. This means that benefits must outweigh the side effects. It is almost impossible to create a drug with no side effects at all. Hence, both the prescription and over the counter (OTC) drugs have adverse effects.1

According to the WHO-UMC definition, ‘a safety signal refers to information on a new or known side effect that may be caused by a medicine and is typically generated from more than a single report of a suspected side effect.’ Generally, more than one report is needed to consider it as a signal. This depends on severity of the adverse drug reaction and accuracy of the information. It is important to note that a signal does not necessarily indicate a direct causal relationship between a side effect and medicine, it mainly helps in stating a hypothesis. Signal detection is the current way to find a reaction. Although, observational studies (post marketing surveillance) and clinical trials are really helpful in finding an ADR(adverse drug reaction), however it has some shortcomings. Number of participants are few for clinical trials, it is difficult to extrapolate the safety of participants after preclinical trials. Patients are not diverse enough, follow up might be challenging at times, and lastly long-term studies are often not possible due to limited availability of resources. To address its shortcomings, Data Mining Algorithms (DMA) is used more frequently nowadays. ^2^

Rucaparib is an anti-cancerous drug approved by USFDA for the treatment of metastatic castrate resistant prostate cancer and BRCA gene mutated ovarian cancer. It belongs to a category of poly ADP ribose polymerase inhibitors. The well reported adverse drugs reactions of rucaparib are as follows:

Rash (27% to 45%), Anaemia (41% to 43%), Neutropenia -Grades 1 to 4 (22%), Thrombocytopenia, (25% to 35%), ALT/SGPT level raised, Aspartate transaminase level above reference range, Asthenia, Dizziness (Up to 20%), Constipation (27% to 39%), Decrease in appetite (23% to 28%), Diarrhoea (20% to 34%), Indigestion (12%), Nausea (52% to 79%), Stomatitis (28%), Taste sense altered (33%), Vomiting (22% to 37%), Dyspnoea (Up to 20%), Nasopharyngitis and Upper respiratory infection ^3^

To evaluate the potential association between rucaparib and the identified signals, disproportionality analysis is done on the spontaneous reports. Another use is to validate a pharmacological hypothesis about the mechanism of occurrence of a particular ADR. Moreover, application of disproportionality analysis could be used to generate signals from post marketing surveillance. ^4^

## Materials and Methods

### 2.1 Data Source

Relevant data was sourced from FAERS database using OpenVigil 2.0 software. FAERS database is a user-friendly system and used mainly for post marketing surveillance. It contains information about adverse events, patient information and errors in medication. Health care professionals (HCPs), drug manufactures all over the world can report the adverse event into the database. Information from FAERS have been used in pharmacovigilance such as signal detection, drug-drug interaction, drug event, idiosyncratic adverse drug reactions.

### 2.2 Study Design

It is a retrospective case – non case study design. Increased prostatic specific antigen, decreased serum magnesium levels, decreased glomerular filtration rate, decreased blood iron and decreased vitamin D were chosen as events and the patients were considered as cases.

### 2.3 Statistical Analysis

The sole purpose of disproportionality analysis is to create a hypothesis of possible causal relationship between drug and a signal. Proportionality analysis was done using PRR (Proportional Reporting Ratio), ROR (Reporting Odds Ratio), chi square and RRR with 95% confidence interval.

#### Proportional Reporting Ratio (PRR)

It is a simple way to measure the strength of blood iron between a risk factor (drug) and a condition (adverse drug reaction). It can be calculated using the formula:

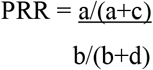

#### Reporting Odds Ratio (ROR)

The odds of occurring of an event with the drug compared to occurrence of same event with all another medicinal product in a database.

It is calculated in a following way,

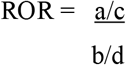

#### Chi square (χ^2^)

Difference between categorical variables from a random sample then a statistical test called **chi square** (χ^2^) is used.

It is calculated by,

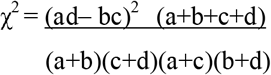

#### Study Procedure

Relevant data was downloaded from OpenVigil 2.1 software^5^ and then the file extracted into the excel software for analysis. Suitable filter was applied per the criteria outlined by Evans et al ^6^ Further this data was selected for analysis.

## Results and Discussion

Results were found to have 21 Drug Events for prostatic specific antigen increased, 13 DEs for-blood magnesium decreased, 10 DEs for glomerular filtration rate decreased, 8 DEs for blood iron decreased and 5 DEs for vitamin D decreased. ROR, RRR and chi square values were obtained as well. A preassigned threshold value was considered and all the values above the threshold value was considered as a positive signal.

During the study period, the FAERS database received a total of 61 reports for increased prostate specific antigen, decreased serum magnesium levels, decreased glomerular filtration rates, blood iron decreased and vitamin D decreased.

In this study we have tried to quantify the risk of increased prostate specific antigen, decreased serum magnesium levels, decreased glomerular filtration rates, blood iron decreased and vitamin D decreased caused by rucaparib with the help of FAERS database which has been queried with OpenVigil 2.1. Upon analysis of the data, we have come across the signal strength of ROR with 10.294, 13.213, 6.951, 6.494 and 4.679 for increased prostate specific antigen, decreased serum magnesium levels, decreased glomerular filtration rates, blood iron decreased and Vitamin d decreased respectively.

### 3.1 Rucaparib

Rucaparib was approved in the year 2016 by USFDA for the treatment of ovarian cancer and in 2020 for prostate cancer. It acts by in inhibiting PARP (poly ADP ribose polymerase). PARPs are the family of proteins which help in repairing the damaged DNA in the cell. Thus, damaging the repair pathways of the DNA and eventually causing cell death. It is indicated for the maintenance treatment of the recurring ovarian cancer and metastatic prostate cancer. ^7^

### 3.2 Prostatic specific antigen increased

Prostatic specific antigen is a protein produced by normal as well as malignant cells of prostate gland. It is a serine protease enzyme which is produced by columnar epithelium of the prostatic gland. The inactive form of PSA (prostatic specific antigen) is pro PSA. Pro PSA gets converted into active PSA while passing through basal and endothelial cell layers of the prostate gland, eventually entering into the circulatory system. Active PSA then binds to protease inhibitors. PSA breaks down the proteins (semenogelin and fibronectin) which is responsible for initial gel like consistency in the semen. This allows the sperm to swim more freely and easily migrate to cervix and promotes fertility.

When prostate gland is in cancerous condition then in fact it reduces the production of PSA and allows the passing of PSA through the basal and endothelial cell layers more easily. Hence its elevated levels in the blood helps in early detection of prostate cancer. As the man ages, the production of spermatozoa is changed causing resistant effect on sperm count and affecting the overall reproductive system. To overcome this, PSA increases as men age and hence promote fertility.^8^

### 3.3 Blood magnesium decreased

Magnesium is present in most of the food items. It acts as a cofactor in more than 300 enzyme systems which regulate the normal metabolism of the body. This electrolyte is required for energy production, glycolysis process and most importantly oxidative phosphorylation. It’s required for the bone strengthening and synthesis of DNA and RNA. Assessing the magnesium status is difficult since magnesium is present inside the cells and bones. The recommended intake of magnesium ranges from 30 mg/day in paediatric population to 51 mg in 50 + years.

Certain drugs including proton pump inhibitors and diuretics may cause decrease in serum magnesium levels. Drugs and magnesium share the same path of metabolism. That means some or the other drug may affect the serum magnesium levels. Consequently, drugs may adversely affect the level of magnesium in the blood. This condition is called hypomagnesemia. It is a common condition and affects 10% of the geriatric population and usually asymptomatic.^9^

### 3.4 Glomerular filtration rate decreased

Glomerular filtration rate represents the flow of plasma into the bowman’s capsule over a specific period of time. About 20% -30% of the cardiac output is received by the kidneys. Out of renal blood flow (RBF) only plasma passes through the membrane of bowman’s capsule. This is called renal plasma flow (RPF) and it is approximately 600-720 ml per minute. Normal glomerular filtration rate (GFR) is 120 ml/min which is 180 litres per day. So, the average urine output becomes 1-2 litres per day. GFR helps determining the pathological condition of the patient mainly the stages of renal failure which ranges from stage 1 to stage 5.

Stage 5 is considered to be kidney failure and the treatment is usually kidney transplant ^10^

Rucaparib which belongs to the category of poly ADP-ribose polymerases inhibitors (PARPi) has been found to promote inflammation process in an ischemic injury. Kidney being very sensitive to the consequences of ischemic injury this could lead to organ damage (kidney). Thus, eventually leading to decreased GFR.^11^

### 3.5 Blood iron decreased

Decreased blood iron levels resulting from chemotherapy are more commonly observed in hematologic, particularly myeloid, malignancies rather than in solid tumors. Among all cancer types, lymphomas, lung tumors, gynecologic, and genitourinary tumors exhibit the highest incidence of anemia, with at least 50%– 60% necessitating transfusions. In the realm of solid tumors, individuals with lung cancer require the highest frequency of transfusions and are typically transfused at higher hemoglobin levels, a phenomenon attributed to both advanced age and suspicion of concurrent pulmonary disease. Prior to commencing chemotherapy, baseline anemia is associated with an increased likelihood of decreased blood iron, and patients with prechemotherapy hemoglobin levels <11 g/dL are more prone to receiving red blood cell transfusions than those with normal baseline hemoglobin levels. Moreover, patients with advanced cancers are generally more anemic at diagnosis and experience poorer survival outcomes ^12^. The extent of decreased blood iron is directly linked to the number of repeated chemotherapy cycles, despite transfusions, indicating limitations in the duration of transfusion benefit.

Decreased blood iron is frequently triggered by platinum-based therapies. Factors associated with the development of platinum-induced anemia include an early decline in hemoglobin post-treatment, cumulative platinum dose, advanced age, chemotherapy non-responsiveness, and a high concentration of residual platinum in the bloodstream after administration. Mechanisms of decreased blood iron by platinum-based regimens encompass the direct suppression of erythroid progenitor cells in the bone marrow and nephrotoxic effects on erythropoietin-producing cells in the kidney. States of inherent erythropoietin deficiency due to cisplatin-induced renal tubular damage can be prevented or treated by replacing the hormone with a recombinant counterpart. Nonplatinum-based chemotherapy regimens, such as antimicrotubular agents, ramapithecines, and biologics, can also be notably myelosuppressive. Fatigue is the predominant symptom of decreased blood iron, although vertigo, loss of appetite, poor concentration, and dyspnea are also commonly reported. The acuity of anemia contributes to symptom severity, with acute-onset anemia resulting in more prominent symptoms. In contrast, progressively developing anemia allows for adaptive mechanisms to compensate for reduced oxygen-carrying capacity, typically without acute symptoms. These adaptive mechanisms include increases in coronary blood flow and cardiac output, as well as alterations in blood viscosity and oxygen utilization^13^.

### 3.6 Vitamin d decreased

Santini and colleagues observed a significant decrease in 25(OH)D levels among breast cancer patients undergoing anthracyclines and taxane anti-tumor treatment, suggesting that nearly all breast cancer patients may experience vitamin D deficiency. A plausible hypothesis is that certain antineoplastic drugs, like taxol, act as ligands for the pregnant X receptor, thereby increasing the catabolism of 25(OH)D and 1,25(OH)2D, resulting in vitamin D deficiency. In cancer patients, vitamin D deficiency is linked to the development of oral mucosa inflammation (mucositis) and taste disturbances (dysgeusia) during chemotherapy. Case studies indicate successful treatment of mucocutaneous side effects (e.g., stomatitis) and dysgeusia in cancer patients undergoing chemotherapy with regimens like TCH (docetaxel, carboplatin, trastuzumab) or FOLFOX6 (fluorouracil, folic acid, oxaliplatin) through vitamin D supplementation. Considering that certain cytostatic agent (e.g., methotrexate) can also have bone-damaging effects and that breast cancer patients often undergo anti-estrogen therapy post-chemotherapy, it is advisable to regularly monitor the vitamin D status in breast cancer patients to gain a comprehensive understanding of the vitamin D levels in these critically ill individuals^14^.

## Conclusion

To avoid the emergence of drug toxicity health care professionals should take extra precaution regarding the aforementioned ADRs. There is growing demand for more drugs and thus there is need for disproportionality analysis. Since this study design helps in forming a hypothesis thus cohort and pharmacoepidemiologic studies are recommended to validate the results found in this study.

## Data Availability

All data produced in the present study are available upon reasonable request to the authors

## Acknowledgment

Our sincere gratitude to the Department of Pharmacy Practice, Faculty of Pharmacy, Ramaiah University of Applied Sciences, for giving us space, time and guidance to conduct our study.

## List of tables

**Table 1.** Shows 2×2 contingency table for disproportionality analysis.

|  | Drug of Interest | Other Drugs | Total |
| --- | --- | --- | --- |
| Event of Interest | a | b | a+b |
| Another event | c | d | c+d |
| Total | a+c | b+d | a+b+c+d |

2×2 contingency table for disproportionality analysis where ‘a’ is the number of reports with the adverse event of interest of the drug of interest, ‘b’ is the number of reports with adverse event of interest for the all-other drugs, ‘c’ is the number of reports for other event of interest for the drug of drug of interest and ‘d’ being the number of reports for the other event of interest for the other drug of interest.

**Table 2.** Shows the association between rucaparib and the identified signals.

| | PRR | $\chi^2$ | ROR | DE |
| --- | --- | --- | --- | --- |
| Increased prostate specific antigen | 10.238 | 165.468 | 10.294 | 21 |
| Decreased serum magnesium levels | 13.167 | 133.699 | 13.213 | 13 |
| Decreased glomerular filtration rates | 6.934 | 44.936 | 6.951 | 10 |
| Blood iron decreased | 6.482 | 31.749 | 6.494 | 8 |
| Vitamin d decreased | 4.674 | 10.983 | 4.679 | 5 |

**Table 3.** Shows the comparison of adverse event of interest for rucaparib vs all other drugs for Prostatic specific antigen increased.

|  | Drug of Interest | All other drugs | Sum total |
| --- | --- | --- | --- |
| Adverse event(s) of interest | 29 | 6573 | 6602 |
| All other adverse event(s) | 4095 | 11144409 | 11148504 |
| Sum total | 4124 | 11150982 | 11155106 |

PRR: 11.93 (8.294; 17.159) RRR: 11.882 (8.261; 17.09) $\chi^2$ : 278.498

**Table 4.** Shows the comparison of adverse event of interest for rucaparib vs all other drugs for Blood magnesium decreased.

|  | Drug of Interest | All other drugs | Sum total |
| --- | --- | --- | --- |
| Adverse event(s) of interest | 20 | 3251 | 3271 |
| All other adverse event(s) | 4104 | 11147731 | 11151835 |
| Sum total | 4124 | 11150982 | 11155106 |

PRR: 16.634 (10.729; 25.791) RRR: 16.539 (10.667; 25.643) $\chi^2$ : 276.83

**Table 5.** Shows the comparison of adverse event of interest for rucaparib vs all other drugs for glomerular filtration rate decreased.

|  | Drug of Interest | All other drugs | Sum total |
| --- | --- | --- | --- |
| Adverse event(s) of interest | 13 | 4658 | 4671 |
| All other adverse event(s) | 4111 | 11146324 | 11150435 |
| Sum total | 4124 | 11150982 | 11155106 |

PRR: 7.546 (4.382; 12.995) ROR: 7.567 (4.387; 13.053) $\chi^2$ : 67.262

**Table 6.**
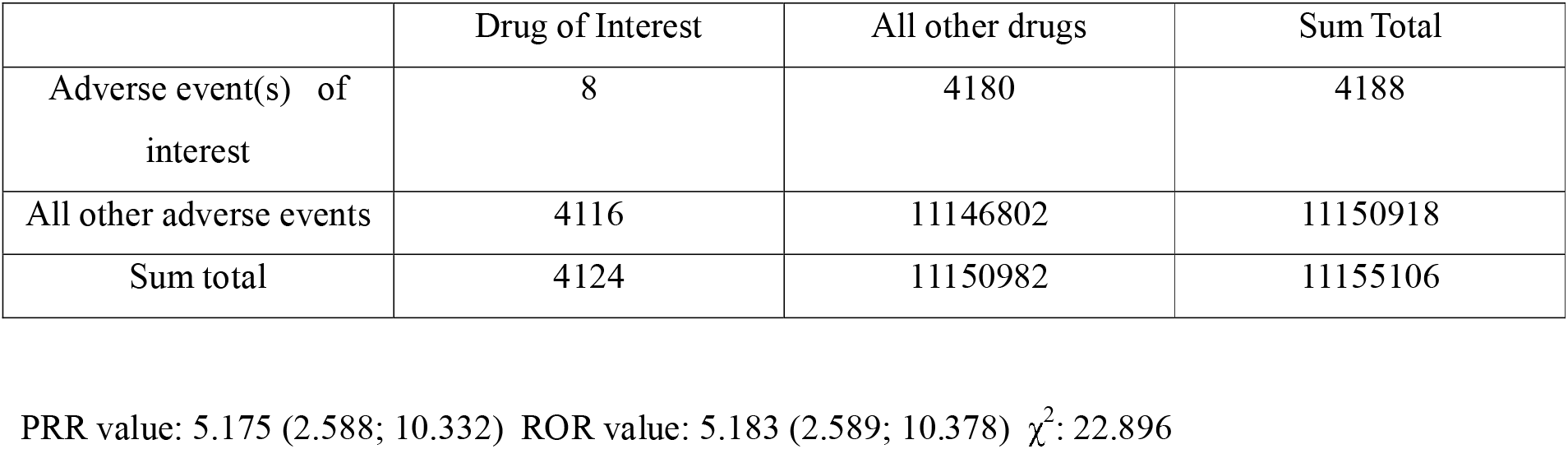
Show the comparison of adverse event of interest for rucaparib vs all other drugs for blood iron decreased.

**Table 7.** Shows the comparison of adverse event of interest for rucaparib vs all other drugs for Vitamin D decreased.

|  | Drug of Interest | All other drugs | Sum total |
| --- | --- | --- | --- |
| Adverse event(s) of interest | 5 | 3461 | 3466 |
| All other adverse event(s) | 4119 | 11147521 | 11151640 |
| Sum total | 4124 | 11150982 | 11155106 |

PRR: 3.906 (1.626; 9.386) ROR: 3.91 (1.625; 9.405) $\chi^2$ : 8.09

## Notes

### Competing Interest Statement

The authors have declared no competing interest.

### Funding Statement

This study did not receive any funding

## References

1. US Food and Drug Administration: Finding and Learning about side effects (adverse reactions). https://www.fda.gov/drugs/find-information-about-drug/finding-and-learning-about-side-effects-adverse-reactions#:~:text=Unwanted%20or%20Unexpected%20Drug%20Reactions%text=Side%20effects%2C%20also%20known%20as (Accessed Aug 8, 2023).

2. Uppsala Monitoring Centre: What is a signal? https://who-umc.org/signal-work/what-is-a-signal/ (Accessed Aug 8, 2023)

3. Micromedex Products https://www.micromedexsolutions.com (Accessed December 9, 2023)

4. Montastruc JL, Sommet A, Bagheri H, Lapeyre-Mestre M. Benefits and strengths of the disproportionality analysis for identification of adverse drug reactions in a pharmacovigilance database. British Journal of Clinical Pharmacology. 2011 Nov 9;72(6):905–8.

5. Böhm R, von Hehn L, Herdegen T, Klein HJ, Bruhn O, Petri H, Höcker J. OpenVigil FDA - Inspection of U.S. American Adverse Drug Events Pharmacovigilance Data and Novel Clinical Applications. PLoS One. 2016 Jun 21;11(6):e0157753. doi: 10.1371/journal.pone.0157753. PMID: 27326858; PMCID: PMC4915658. https://pubmed.ncbi.nlm.nih.gov/27326858/

6. Evans SJ, Waller PC, Davis S. Use of proportional reporting ratios (PRRs) for signal generation from spontaneous adverse drug reaction reports. Pharmacoepidemiol Drug Saf. 2001 Oct-Nov;10(6):483–6. doi: 10.1002/pds.677. PMID: 11828828. https://pubmed.ncbi.nlm.nih.gov/11828828/

7. Research C for DE and. Rucaparib. FDA. https://www.fda.gov/drugs/resources-information-approved-drugs/rucaparib (Accessed January 9, 2024)

8. Prostate-Specific Antigen (PSA) Test -National Cancer Institute. https://www.cancer.gov. 2017. https://www.cancer.gov/types/prostate/psa-fact-sheet#:~:text=Prostate%2Dspecific%20antigen%2C%20or%20PSA (Accessed February 3, 2024)

9. National Institutes of Health. Office of Dietary Supplements - Magnesium. National Institutes of Health. 2016. https://ods.od.nih.gov/factsheets/Magnesium-HealthProfessional/ (Accessed March 7, 2024)

10. Kaufman DP, Basit H, Knohl SJ. Physiology, Glomerular Filtration Rate (GFR). National Institute of health StatPearls Publishing; 2019. https://www.ncbi.nlm.nih.gov/books/NBK500032/ (Accessed March 20, 2019)

11. Deshpande PP, Perazella MA, Jhaveri KD. PARP inhibitors and the Kidney. Journal of Onco-Nephrology. 2021;5(1):42–47

12. Spivak JL, Pere Gascón, Ludwig H. Anemia Management in Oncology and Hematology. Oncologist. 2009 Sep 1;14(S1):43–56 https://pubmed.ncbi.nlm.nih.gov/19762516/

13. Rothmann SA, Paul P, Weick JK, McIntyre WR, Fantelli F. Effect of cis-diamminedichloroplatinum on erythropoietin production and hematopoietic progenitor cells. International Journal of Cell Cloning. 1985;3(6):415–23. Available from: https://pubmed.ncbi.nlm.nih.gov/4067362/

14. Wood PA, Hrushesky WJ. Cisplatin-associated anemia: an erythropoietin deficiency syndrome. J Clin Invest. 1995 Apr;95(4):1650–9. https://pubmed.ncbi.nlm.nih.gov/7706473/

